# Racial/Ethnic Disparities Impact the Real-World Effectiveness of a Multicomponent Maternal Smoking Cessation Program: Findings from the CTTP Cohort

**DOI:** 10.1101/2023.06.26.23291787

**Authors:** Stacey D. Wiles, Jerry W. Lee, Anna Nelson, Anne Berit Petersen, Pramil N. Singh

**Author notes:** Corresponding Author: Dr. Anne Berit Petersen.

## Abstract

**Introduction:** Smoking during pregnancy adversely affects perinatal outcomes for both women and infants. We conducted a retrospective cohort study of the state-funded Comprehensive Tobacco Treatment Program (CTTP) – the largest maternal tobacco cessation program in San Bernardino County, California – to determine the real-world program effectiveness and to identify variables that can potentially improve effectiveness.

**Methods:** During 2012-2019, women who smoked during pregnancy were enrolled in CTTP’s multicomponent behavioral smoking cessation program that implemented components of known efficacy (i.e., incentives, biomarker testing, feedback, and motivational interviewing).

**Results:** We found that 40.1% achieved prolonged abstinence by achieving weekly, cotinine-verified, 7-day abstinence during 6 to 8 weeks of enrollment. Using intention-to-treat analyses, we computed that the self-reported point prevalence abstinence rate (PPA) at the six-month telephone follow-up was 36.7%. Cohort members achieving prolonged abstinence during the CTTP were five times more likely to achieve PPA six months after CTTP. Several non-Hispanic ethnicities (Black, Native American, White, or More than one ethnicity) in the cohort were two-fold less likely (relative to Hispanics) to achieve prolonged abstinence during CTTP or PPA at six months after CTTP. This disparity was further investigated in mediation analysis. Variables such as quitting during the first trimester and smoking fewer cigarettes at enrollment were also associated with achieving PPA at six months.

**Discussion:** Racial/ethnic health disparities that have long been linked to a higher rate of maternal smoking persist even when the pregnant smoker enrolls in a smoking cessation program.

**Significance:** *What is already known on this subject?*

Causal factors that enable pregnant smokers to achieve abstinence during and after enrollment in maternal smoking cessation programs are not well understood.

*What does this study add?*

We found that US (United States) racial/ethnic disparities that have long been linked to higher national rates of maternal smoking for several non-Hispanic ethnicities (Black, Native American/Alaskan, White) also persisted in the form of lower abstinence rates in non-Hispanic ethnicities (Black, Native American, White, More than one ethnicity) who enrolled in the maternal smoking cessation program we evaluated. This racial/ethnic disparity in maternal smoking cessation program outcomes is a new addition to the literature and needs further study to identify the underlying causal factors.

## Introduction

US birth certificate data from 2021 indicates that the prevalence of women who smoked cigarettes at any time during their pregnancy was 4.6% (Martin, J.K.A., & Driscoll, 2023). In San Bernardino County, California, the largest county in the contiguous United States, birth certificate data indicate that there are approximately 1,300 pregnancies every year among women who smoke (Batech et al., 2013; “California Births by County and Month, January 2018 to November 2020,” 2020). Maternal smoking increases the risks of placental previa and abruption, miscarriage, stillbirth, fetal growth restriction, preterm delivery, low birth weight, and neonatal mortality (Abraham et al., 2017; Banderali et al., 2015; Berlin, Golmard, Jacob, Tanguy, & Heishman, 2017; Liu et al., 2020; Pineles, Park, & Samet, 2014; Shobeiri & Jenabi, 2017; Shobeiri, Masoumi, & Jenabi, 2017). In the US, maternal smoking trends indicate a strong racial/ethnic disparity. Specifically, maternal smoking rates are highest in Native American, Non-Hispanic White, Non-Hispanic Black, and some Pacific Islander women, and lowest in Hispanic and Asian women. Also, women with lower socioeconomic status, Medicaid insurance, and WIC enrollment have higher odds of smoking and lower odds of quitting during pregnancy (Curtin & Matthews, 2016).

The Comprehensive Tobacco Treatment Program (CTTP) is a state-funded (First5CA.gov), multicomponent (i.e., incentives, biomarker testing, feedback, and motivational interviewing) behavioral smoking cessation program that was designed and implemented by Loma Linda University Health (LLUH) from 2012 to 2019 to reduce smoking-related morbidity and mortality among pregnant smokers and their children in San Bernardino County, California. As part of the program evaluation of CTTP, the data from all CTTP enrollees were analyzed as a retrospective cohort study that has been previously described (Petersen et al., 2021). The objectives of this retrospective cohort study based on the CTTP data are (1) to determine the real-world effectiveness of a multicomponent cessation program on smoking during pregnancy and on follow-up, (2) to determine what maternal demographic predictors influence continued smoking during pregnancy and self-reported smoking six months after program participation.

## Methods

### CTTP Cohort

The CTTP cohort has been comprehensively described in a cohort profile report by Petersen et al (2021). The cohort consists of 1,402 women in San Bernardino County who smoked during pregnancy and enrolled in the CTTP between 2012 and 2019. Briefly, the cohort was enrolled in a 6 to 8 week multicomponent behavioral smoking cessation program. The goal of the program was to achieve prolonged abstinence as defined by weekly cotinine-verified (urinary cotinine) point-prevalence abstinence being achieved each week of enrollment. During 2012-2019, program staff consistently collected self-reported point prevalence abstinence at six months by conducting a telephone follow-up.

### CTTP Multicomponent Behavioral intervention

The intervention components were extensively described in a cohort profile report (Petersen et al., (2021). Briefly, the intervention consisted of an individual intake appointment to work with trained health educators to develop an individualized quit plan that involved setting a quit date and completing the following components:

1. The 5 A’s counseling and motivational interviewing technique using evidence-based materials developed by the American College of Obstetricians and Gynecologists,
2. feedback on smoking-related exposures and harms,
3. weekly urinary cotinine testing and discussion of the results with the cohort members, and
4. incentives (infant diapers and xylitol gum to prevent or reduce cravings) were provided each week to cohort members testing negative for cotinine.

The weekly counseling sessions were done in a group classroom setting and lasted about 1 hour. Cohort member records and databases were created, entered, and maintained under confidential patient data conditions by the CTTP health educators.

### Statistical Methods

The real-world effectiveness of CTTP was assessed by computing prolonged abstinence (PA) as being the proportion and 95% confidence interval of cohort members who achieved weekly cotinine-verified 7-day abstinence during enrollment in CTTP. During 2012-2017, the criteria for PA was eight consecutive weeks of cotinine-verified abstinence. During 2017-2019, the criteria was changed to six consecutive weeks. CTTP effectiveness was also assessed by computing self-reported (during telephone follow-up) point prevalence abstinence (PPA) and 95% confidence interval (CI) at six months. PA and PPA were computed for the whole cohort and for subgroups by PA criteria (eight weeks versus six weeks).

Variables associated with the binary PA and Six-month PPA outcomes were assessed in logistic regression models that tested demographic (age, race/ethnicity), health (parity, trimester at enrollment), and nicotine dependence (number of cigarettes smoked at enrollment) predictor variables. Odds ratios and 95% CI were computed for these variables. Effectiveness was also tested in logistic regression models with 6-month PPA as the outcome and PA as the predictor variable. Preliminary contingency table analysis and Pearson chi-squared tests were performed relating the outcome variables to the predictor variables. Confounding scenarios were tested using a change of estimate approach. Models were run in the whole cohort and for subgroups by PA criteria (eight weeks versus six weeks).

The Hayes PROCESS macro for SPSS—SPSS Statistics 27 (IBM, 2020)—was used to assess mediating factors in the relationship between ethnicity and abstinence at program end (Hayes, 2017). Cohort Members were categorized into those who were enrolled under the eight-week case definition for having quit (program years July to June 2012-2017) and those enrolled under the six-week case definition (program years July to June 2017-2019). PA during the program and PPA at six months were compared between cohort members who were enrolled under the eight-week PA criteria and those enrolled under the six-week PA criteria. Loma Linda University Health Institutional Review Board granted approval to analyze individually de-identified data.

Missing outcome data for PA and PPA were assumed to be not abstinent per an intention-to-treat protocol (White et al., 2011). The proportion with missing data for variables added to the multivariable models was < 1% per variable and was handled for the main analysis by exclusion. A sensitivity analysis with multiple imputations on the predictor variables indicated no difference in the point estimates or confidence intervals in the models. Modeling of the datasets was done in SPSS Statistics 27 (IBM, 2020) and in SAS version 9.4 (Cary, NC).

## Results

The basic demographic, behavioral, and gestational profile of the CTTP cohort has been previously reported (Petersen et al., 2021) and indicates that the cohort has a mean age of 27 years at enrollment and was 43% Hispanic.

### Real-world Effectiveness of CTTP

We found that 40.1%, 95% CI [37.5, 42.7], of cohort members achieved PA by program end. In Table 1, we computed PA for levels of pertinent demographic and health variables and found significantly higher level of PA among those cohort members who were older age, Hispanic ethnicity, enrolled during the second trimester, and smoked fewer cigarettes at enrollment. By PA criteria, we found that the stricter criteria of achieving abstinence for eight weeks had a lower level of PA (37.0%, 95% CI [34.0, 44.0]) than the six-week criteria (47%, 95% CI [42.3, 51.5]). This may be artifactual due to the stricter criteria of including two more weeks of cotinine-verified surveillance for PA assessment in the “eight week” group.

**Table 1.** Prolonged abstinence by the end of the program^a^ is given by level of pertinent demographic and health variables reported by the Comprehensive Tobacco Treatment Program for the 2012-2019 cohort (*N* = 1,402).

| **Demographic and Health Variables** | **Prolonged Abstinence (%)** | **95% Confidence Interval** |  |
| --- | --- | --- | --- |
| **Maternal age in years\*\*** |  |  |  |
| 20 years or younger | 29.5% | 23.0% | 36.0% |
| 21-25 years | 34.8% | 30.4% | 39.3% |
| 26-30 years | 41.3% | 36.4% | 46.3% |
| 31 years or older | 50.4% | 45.4% | 55.4% |
| Missing |  |  |  |
| **Maternal race/ethnicity\*\*** |  |  |  |
| Hispanic | 54.0% | 50.0% | 58.0% |
| Non-Hispanic White | 27.0% | 22.8% | 31.3% |
| Non-Hispanic Black | 31.6% | 26.1% | 37.1% |
| Asian/Pacific Islander | 50.0% | 29.1% | 70.9% |
| Native American | 27.3% | 1.0% | 53.6% |
| More than one ethnicity | 31.4% | 20.6% | 42.3% |
| **Trimester at enrollment\*** |  |  |  |
| First | 32.4% | 26.8% | 37.9% |
| Second | 46.0% | 41.5% | 50.5% |
| Third | 39.1% | 35.3% | 42.8% |
| **Number of Cigarettes Smoked\*\*** |  |  |  |
| None | 51.4% | 48.2% | 54.6% |
| 5 or fewer | 19.1% | 14.7% | 23.5% |
| More than 5 | 11.3% | 6.2% | 16.3% |
| **Number of pregnancies** |  |  |  |
| 1 | 41.1% | 35.8% | 46.4% |
| 2 | 39.3% | 33.7% | 44.9% |
| 3 | 41.5% | 35.1% | 48.0% |
| 4 | 44.9% | 38.1% | 51.7% |
| 5 or more | 36.8% | 31.7% | 41.9% |

<sup>a</sup>Achieved cotinine verified 7-day abstinence each week of the 6-8 week CTPP intervention.
\* $p < 0.005$ for chi-squared test for independence between prolonged abstinence and demographic/health variable.
\*\* $p < 0.001$ for chi-squared test for independence between prolonged abstinence and demographic/health variable.

At the six-month follow-up after the program, 36.7%, 95% CI [34.2, 39.3], of cohort members reported seven-day PPA. We found no difference in 6-month PPA between the group 8-week PA criteria group (36.2%, 95% CI [31.7, 40.6]) and the six-week PA criteria group (37.0%, 95% CI [33.9, 40.0]).

### Variables associated with Cotinine-verified PA during CTTP Enrollment

In univariable logistic regression models, we found increased odds of PA for older age (OR [95% CI] per 1 year increment = 1.05 [1.03,1.07]) and decreased odds of PA for number of cigarettes smoked at enrollment (OR [95% CI] per cigarette = 0.75 [0.70, 0.79]), non-Hispanic ethnicity (OR [95% CI] relative to Hispanic = 0.39 [0.29, 0.529] for non-Hispanic Black; 0.31 [0.24, 0.41] for non-Hispanic white; 0.32 [0.08, 1.21] for Native American); 0.85 [0.36, 1.98] for Asian/Pacific Islander; 0.37 [0.22, 0.63] for More than one ethnicity) and early and late gestation at enrollment (OR [95% CI] relative to trimester 2 = 0.56 [0.41, 0.77] for trimester 1; 0.75 [0.59, 0.96] for trimester 3). We also found increased odds of PA for the less strict 6-week criteria for PA (OR relative to 8 weeks = 1.50 [1.20, 1.89]). We found no important association between PA and the number of pregnancies.

In Table 2, we added pertinent demographic and health variables that showed strong univariable associations with PA to a multivariable model. In this multivariable model, associations seen in univariable models remained. We found that non-Hispanic ethnicities (non-Hispanic Black, non-Hispanic White, Native American, More than one ethnicity) trended towards a more than two-fold decrease in odds of achieving PA by program end. Older age, fewer cigarettes smoked at enrollment, second trimester at enrollment, and less strict criteria for PA (six versus eight weeks of cotinine-verified abstinence) were all associated with higher odds of achieving PA.

**Table 2.** Multivariable logistic regression model relating prolonged abstinence by the end of the program^a^ to pertinent demographic and health variables reported by the Comprehensive Tobacco Treatment Program for the 2012-2019 Cohort (*N* = 1,402).

| Demographic and Health Variables | OR | 95% Confidence Interval |
| --- | --- | --- |
| Enrollment age (per 1 year increment) | 1.06 | 1.04 to 1.09 |
| Number of Cigarettes smoked at enrollment (per 1 cigarette increment) | 0.75 | 0.71 to 0.81 |
| Race/Ethnicity |  |  |
| Hispanic | 1 | [referent] |
| non-Hispanic Black | 0.44 | 0.32 to 0.60 |
| non-Hispanic White | 0.45 | 0.33 to 0.60 |
| Native American | 0.36 | 0.09 to 1.47 |
| Asian/Pacific Islander | 0.84 | 0.34 to 2.04 |
| More than one ethnicity | 0.39 | 0.22 to 0.69 |
| Trimester at Enrollment |  |  |
| trimester1 | 0.61 | 0.43 to 0.86 |
| trimester 2 | 1 | [referent] |
| trimester3 | 0.59 | 0.45 to 0.77 |
| Program Criteria for Prolonged Abstinence |  |  |
| 6 week prolonged Abstinence | 1.51 | 1.18 to 1.94 |
| 8 week prolonged Abstinence | 1 | [referent] |

<sup>a</sup>Achieved cotinine verified 7-day abstinence each week of the 6-8 week CTPP intervention.

In Table 3, we conducted a mediation analysis to identify possible mediators of the effect where a higher proportion achieved PA among those with Hispanic ethnicity (Table 2). We found evidence for partial mediation whereby the higher proportion achieving PA among Hispanic cohort members was mediated through their smoking fewer cigarettes at enrollment and, to a lesser extent, older age at enrollment. Weeks of gestation had an inverse parabolic effect whereby enrollment during trimester 2 had the highest PA. Weeks of gestation did not seem to mediate the Hispanic ethnicity effect and Figure 1 shows that Hispanics had a higher PA than non-Hispanics during CTTP enrollments occurring throughout gestation.

**Figure 1.**
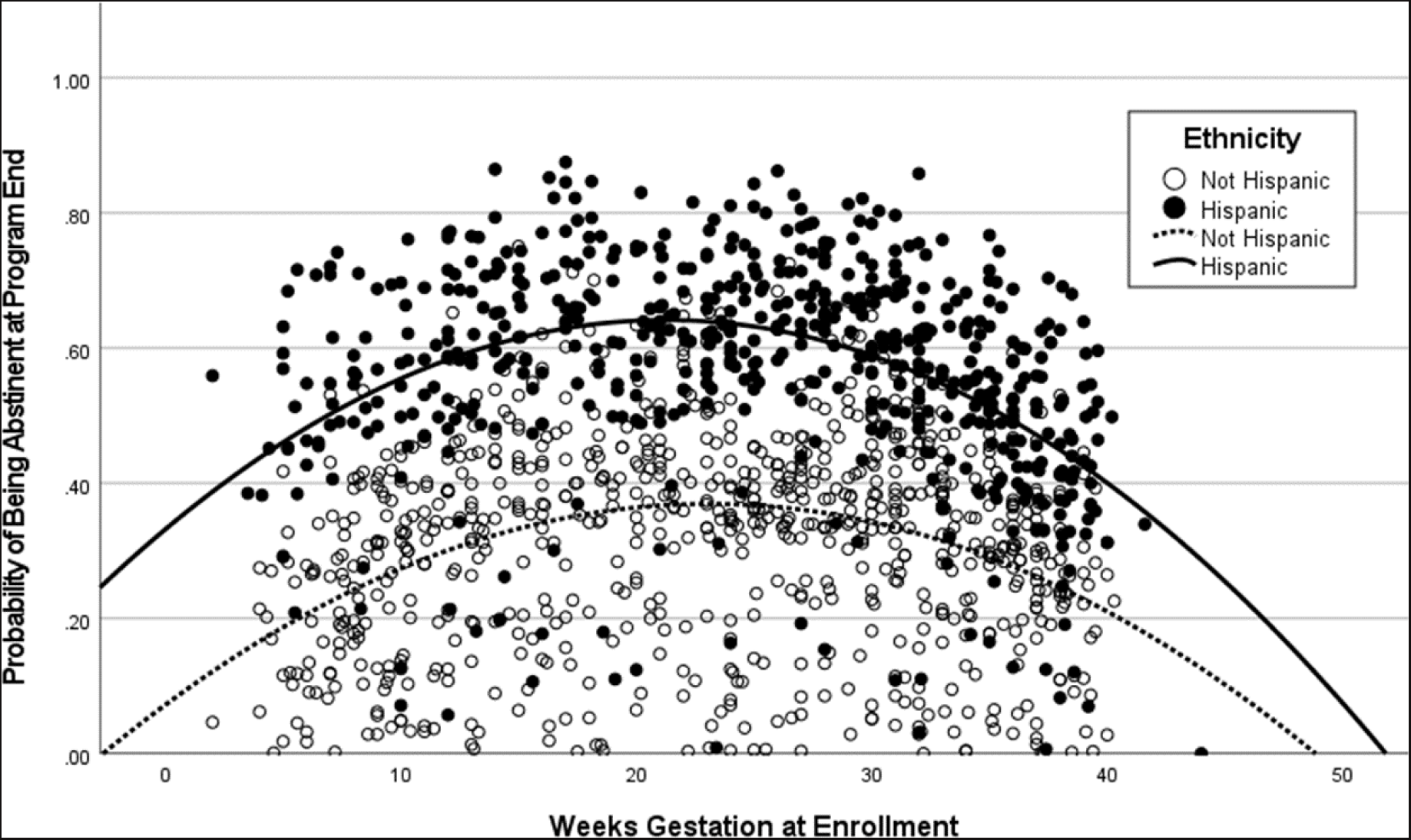
Relationship of weeks gestation at enrollment and probability of abstinence at program end by ethnicity for cohort members in the Comprehensive Tobacco Treatment Program, 2012-2019, (*N* = 1,402).

**Table 3.** Multivariable logistic regression model investigating mediators^a^ of the positive relation between Hispanic ethnicity and achieving prolonged abstinence in the 2012-2019 cohort of the Comprehensive Tobacco Treatment Program (*N* = 1,402).

| **Variable** | **Odds of Prolonged Abstinence**<br>**at Program End**<br>**(95% CI)** |
| --- | --- |
| **Variable Effect on Prolonged Abstinence** |  |
| Hispanic ethnicity | 2.23 (1.74-2.85)*** |
| Maternal age | 1.09 (1.06-1.11)*** |
| Weeks gestation at enrollment | 1.13 (1.06-1.20)*** |
| Weeks gestation at enrollment, squared term | <1.00 (<1.00-<1.00)*** |
| Number of cigarettes per day at enrollment | 0.77 (0.73-0.83)*** |
| Number of pregnancies | 0.86 (0.78-0.94)** |
| Constant | 0.05 (0.02-0.11)*** |
| **Direct Effect of Hispanic Ethnicity** | 2.23 (1.74-2.85)*** |
| **Indirect Effect of Hispanic Ethnicity** |  |
| Maternal age | 1.07 (1.01-1.14)* |
| Weeks gestation at enrollment | 1.29 (1.10-1.60)* |
| Weeks gestation at enrollment, squared term | 0.76 (0.60-0.89)* |
| Number of cigarettes per day at enrollment | 1.52 (1.30-1.92)* |
| Number of pregnancies | 1.00 (0.97-1.03) |
| Total | 1.58 (1.34-2.01)* |
| **Total Effect of Hispanic Ethnicity** | 2.75 (2.19-3.44)* |

<sup>a</sup>Analyses used Hayes PROCESS macro for SPSS with 5000 bootstrap samples.
\**p* ≤ .05
\*\**p* ≤ .01
\*\*\**p* ≤ .001

### Variables associated with Self-reported PPA at Six months of follow-up after CTTP participation

In univariable logistic regression models, the strongest effect found was that those cohort members who achieved PA during CTTP enrollment were five-fold more likely (relative to those who did not) to achieve PPA (OR [95% CI] = 5.51 [4.35, 6.98]) six months after CTTP participation. This effect remains for cohort members achieving PA through the eight-week criteria (OR [95% CI] = 6.31 [4.72, 8.45]) or six-week criteria (OR [95% CI] = 4.49 [2.97, 6.81]).

We also found increased odds of achieving PPA at six months for older age (OR [95% CI] per 1 year increment = 1.02 [1.01, 1.04]) and decreased odds of PA for the number of cigarettes smoked at enrollment (OR [95% CI] per cigarette = 0.83 [0.79, 0.87]), non-Hispanic ethnicity (OR [95% CI] relative to Hispanic = 0.48 [0.36, 0.65] for non-Hispanic Black; 0.36 [0.27, 0.47] for non-Hispanic white; 0.90 [0.27, 2.97] for Native American); 0.75 [0.31, 1.77] for Asian/Pacific Islander; 0.47 [0.28, 0.81] for More than one ethnicity). We found no association with trimester at enrollment (OR [95% CI] relative to trimester 2 = 1.04 [0.76, 1.42] for trimester 1; 1.08 [0.84, 1.38] for trimester 3), the less strict eight-week criteria for PA (OR relative to eight weeks = 0.97 [0.77, 1.22]), or the number of pregnancies (OR per pregnancy = 1.00 [0.93, 1.06]). Due to finding no difference in PA criteria, we did not add the variable to the final model and combined the six-week and eight-week cohorts for a whole cohort analysis.

In Table 4, we added pertinent demographic and health variables that showed strong univariable associations with PPA at 6 months to a final multivariable model. We found significantly higher odds of PPA at six months for those cohort members who attained PA during CTTP, older age at enrollment, number of cigarettes smoked at enrollment, and enrolling during the first trimester.

**Table 4.** Multivariable logistic regression model relating point prevalence abstinence at six months follow-up after the program to pertinent demographic and health variables reported by the Comprehensive Tobacco Treatment Program for the 2012-2019 Cohort (*N* = 1,402).

| Demographic and Health Variables | OR | 95% Confidence Interval |
| --- | --- | --- |
| **Enrollment age (per 1 year increment)** | 1.01 | 0.99 to 1.03 |
| **Number of Cigarettes smoked at enrollment (per 1 cigarette increment)** | 0.91 | 0.87 to 0.96 |
| **Race/Ethnicity** |  |  |
| Hispanic | 1 | [referent] |
| non-Hispanic Black | 0.67 | 0.48 to 0.93 |
| non-Hispanic White | 0.56 | 0.41 to 0.76 |
| Native American | 1.43 | 0.39 to 5.18 |
| Asian/Pacific Islander | 0.79 | 0.31 to 2.00 |
| More than one ethnicity | 0.65 | 0.36 to 1.16 |
| **Trimester at Enrollment** |  |  |
| Trimester 1 | 1.45 | 1.02 to 2.06 |
| Trimester 2 | 1 | [referent] |
| Trimester 3 | 1.14 | 0.86 to 1.50 |
| **Prolonged Abstinence at Program End** |  |  |
| Yes | 4.46 | 3.46 to 5.75 |
| No | 1 | [referent] |

## Discussion

Our findings from a multi-ethnic cohort of pregnant smokers living in San Bernardino County, California, identified the real-world effectiveness of a multicomponent behavioral smoking cessation program using components of known efficacy and trained health educators. In the CTTP cohort, we found that:

1. 40.1% of the cohort achieved PA from smoked cigarettes by the end of CTTP by achieving weekly, cotinine-verified 7-day abstinence during the 6-8 week intervention phase,
2. 36.7% of the cohort achieved self-reported PPA at six months after CTTP participation,
3. Achieving PA (relative to those who did not) during the intervention phase was associated with a significant 5-fold increase in odds (OR [95% CI] = 5.51 [4.35, 6.98]) of achieving PPA at six months post-intervention, and
4. some of the non-Hispanic ethnicities (Black, Native American, White, More than one ethnicity) in the cohort were 2-fold less likely to achieve PA during CTTP or PPA at six months after CTTP.

Other variables associated with significantly higher rates of abstinence at 6 months after CTTP include enrollment during the first trimester, fewer cigarettes smoked at enrollment, and older age at enrollment.

### Racial/ethnic disparities in Maternal Smoking and Maternal Smoking Cessation

In the CTTP cohort, we found that Hispanic cohort members achieved significantly higher odds of abstinence during and six months after the intervention. In contrast, the lowest abstinence levels during and after the intervention were found for cohort members who were non-Hispanic Native American, non-Hispanic white, non-Hispanic Black, or had reported more than one ethnicity. These trends closely follow 2016-2021 national trends in maternal smoking, where the highest rates of current smoking during pregnancy are observed among non-Hispanic Native Americans, non-Hispanic whites, and non-Hispanic blacks (2023). Our cohort did not have the power to detect the significantly higher rates of maternal smoking in Native American and certain Pacific Islander (i.e., Samoan) communities that are seen in national and California state data sets(Hoshiko et al., 2019; Martin et al., 2023).

Thus, our findings show that health disparities based on race/ethnicity that produce a higher rate of maternal smoking for Native American, non-Hispanic Black, and non-Hispanic whites persist even when the pregnant smoker enrolls in a smoking cessation program and attempts to achieve short-term or long-term abstinence. To our knowledge, we are the first study to show that the disparity continues after enrollment in maternal smoking cessation. National data (NHANES, US Birth Certificate) show that overall maternal smoking cessation rates (i.e., self-directed cessation, enrollment in a cessation program) are highest in Hispanic and lowest in non-Hispanic Black and non-Hispanic White (Curtin & Matthews, 2016; Li et al., 2018). The factors underlying this racial/ethnic disparity are not well understood, and identifying these factors is key to improving the effectiveness of US maternal smoking cessation programs.

In the CTTP cohort, our mediation analysis of the higher proportion achieving abstinence among Hispanics identified that some of this effect may be partially mediated by their lower smoking rate, enrollment at the second trimester, and older age at the time of enrollment in maternal smoking cessation. These factors provide an incomplete picture of the mediation, and since CTTP was not formally designed as a behavioral study, we do not have variables to further investigate effects.

Potential mediators and pathways from other studies are discussed here. One factor could be the Hispanic Paradox that posits that culture-based social support and less acculturation in the community overcomes health disparities typically caused by poverty (Fishman, Morgan, & Hummer, 2018). Among Mexican women, Noah et al. has reported that their residence in a Hispanic-majority community was associated with lower maternal smoking rates as compared to those residing in a non-Hispanic community (Noah, Landale, & Sparks, 2015). We found evidence of partial mediation of the Hispanic-abstinence relation by an older age-abstinence pathway. Some of this effect with older age could be related to its association with less acculturation (Table 3).

Another possible pathway underlying the racial/ethnic difference we observed in abstinence achieved in the CTTP cohort are racial/ethnic differences in nicotine metabolism. Jain has reported that the urinary nicotine metabolite ratio follows trends whereby non-Hispanic Black > non-Hispanic White > Hispanic (Jain, 2020). The slower metabolism in Hispanics might explain their smoking fewer cigarettes at CTTP enrollment, with this mechanism potentially underlying the partial mediation of their higher odds of abstinence (Table 3).

When considering the higher proportion achieving abstinence among Hispanic cohort members (42.9% of the cohort), another possible explanation is the concordance in ethnicity these cohort members shared with the three health educators who were Hispanic and Spanish speaking. To address the overall racial disparity evidenced by a 3-4 fold increase in maternal morbidity and mortality in US Black women, Liese et al. developed a multicomponent intervention model that involved racial concordance between all care providers (including patient navigation and health workers support) and patients, group prenatal care, perinatal nurse navigation, and 12 months of in-home postpartum doula support (Liese et al., 2022). In the Aboriginal/Torres Strait population of Australia, indigenous community health workers were shown to be effective in promoting maternal smoking cessation (Mersha et al., 2022) and adult smoking cessation (Kennedy et al., 2022). Research by Cheng and colleagues has demonstrated positive associations between patient-provider racial/ethnic concordance, better working alliance, and longer length of treatment (Cheng, Nakash, Cruz-Gonzalez, Fillbrunn, & Alegría, 2021).

### How effective are maternal smoking cessation programs?

A Cochrane systematic review of over 97 behavioral smoking interventions on 26,637 women who smoked during pregnancy found that enrolling in behavioral smoking cessation programs produced a significant 35% increase in achieving late pregnancy abstinence (Chamberlain et al., 2017). The 2021 United States Preventive Services Taskforce report cited the most commonly used behavioral intervention components as being: counseling (i.e., motivational interviewing), incentives, health education, and social support (Patnode et al., 2021). A systematic review of smoking cessation outcomes after behavioral intervention found counseling and incentives more effective than health education or social support (Chamberlain et al., 2017). Overall, enhanced efficacy has been found with multicomponent interventions that combine an effective component like financial incentives (Boyd, Briggs, Bauld, Sinclair, & Tappin, 2016; Boyd, Tappin, & Bauld, 2016) with other components (Chamberlain et al., 2017) such as:

1. a higher frequency of counseling sessions,
2. tailoring of messaging and materials to pregnant persons,
3. messaging that addresses both maternal and fetal health effects of smoking, and
4. emphasis on quitting early in pregnancy.

Also notable is that an evaluation of the efficacy of the UK’s National “Stop Smoking Services” for pregnant smokers identified the effectiveness of 1) home visits (Bauld et al., 2017), and 2) regular biochemical testing on tobacco use at prenatal visits (Campbell et al., 2017; Sloan et al., 2016).

### Does quitting early in pregnancy increase chances of long-term abstinence?

Our findings from the CTTP cohort indicating higher odds of abstinence at six months after the program for women who enrolled in smoking cessation during the first trimester or at a lower smoking intensity level (Table 4) are supported by data from a few other cohorts (Cooper et al., 2017; Schneider, Huy, Schütz, & Diehl, 2010). A longitudinal study of pregnant women from the UK found a temporal effect whereby quit attempts, reports of intention to quit, and achieving abstinence tend to decrease over time during the period from gestation through 2 months postpartum (Cooper et al., 2017). Interestingly cessation during early pregnancy may also be associated with lower risk of adverse birth outcomes (Yan & Groothuis, 2015).

Despite the concordance of our 6-month follow-up findings with the literature we note the unexpected curvilinear association (Table 2, Figure 1) in which abstinence at the end of the 6-8 week program appears to be highest for those cohort members who enrolled during the second trimester. This unexplained association could be due to non-causal pathways. For example, second trimester enrollees may have started a self-directed smoking reduction or cessation during their first trimester.

### Limitations

Our retrospective cohort study of CTTP effectiveness has some important limitations. The criteria for achieving PA changed from 8 weeks to 6 weeks of cotinine-verified abstinence during the final two years of the program. We modeled the effect of this change and found that it had no effect on PPA at six months (OR [95% CI] for 6 versus 8 weeks = 0.97 [0.77, 1.22]). Thus, most of our analyses were done on the whole cohort. Also, PA achieved through either 6 weeks or 8 weeks of cotinine tests resulted in a substantial increase in the odds of PPA at six months of four-fold and six-fold, respectively. This finding is consistent with USPTF findings that at least four counseling sessions is optimal(Patnode et al., 2021).

As noted in a previous CTTP cohort profile paper (Petersen et al., 2021), six months after the program, more than half the cohort members were lost to follow-up. Since CTTP is a behavior change program and not a randomized control trial there was not infrastructure to maximize subject retention for the sole purpose of data collection. This was also our rationale for reporting these findings as “real world” effectiveness of implementing several intervention components of known efficacy.

When considering our mediation analysis in Table 3, we note that CTTP was designed as a service program and not as a behavioral intervention study. Thus, we do not have the range of behavioral and health variables to fully study mediation. The partial mediation we found needs further investigation in studies designed to find mediators of the racial/ethnic disparity in maternal smoking and achieving abstinence in a maternal smoking cessation program.

We note that, like many smoking cessation studies, we have used only self-report for the assessment at six months. The loss to follow-up rate would have been higher if we had asked cohort members to return for cotinine verification of their abstinence. Prior research shows that self-report of smoking tends to under-report smoking, particularly at low levels of cigarette smoking (England et al., 2007).

Lastly, CTTP did not have linkage with electronic health records or the prenatal care data of the cohort members. Thus, we could not include in the analysis variables pertaining to prenatal health conditions.

## Conclusions

In a multicomponent smoking cessation program, we found the following variables to be associated with abstinence six months after the program: Hispanic ethnicity, older age at enrollment, enrollment during the first trimester, and lower intensity of smoking at enrollment. We found that the highest rates of dropout/relapse from the program were among women who were non-Hispanic Black, non-Hispanic White, Native American/Alaskan Native, or reported More than one ethnicity. This disparity follows national trends in maternal smoking. We conclude from our evaluation that Racial/ethnic health disparities that have long been associated with higher rates of maternal smoking in the US can persist even when the pregnant smoker enrolls in a smoking cessation program.

## Declarations

### Funding

Data collection was funded by First 5 CA.gov (San Bernardino). Initial data analysis and reporting by PS was funded by Grant HHSN 267200700021C from NICHD/Department of Health and Human Services (National Children’s Study Award to University of California, Irvine, and sub-award to Loma Linda University and California State University San Bernardino). ABP and PS conducted further analysis of the CTTP cohort for this report under a Tobacco-related Disease Research Program award T32KT4784 from the University of California.

### Conflicts of Interest

The authors declare that they have no conflict of interest.

### Ethics Approval

The study was conducted according to the guidelines of the Declaration of Helsinki and approved by the Institutional Review Board of Loma Linda University (IRB # 5190418; October 25, 2019).

### Consent to Participate

Patient consent was waived as this was a program evaluation, and no private, individually identifiable information was obtained or received.

### Consent for Publication

Non-applicable.

### Availability of Data and Material

Available upon reasonable request.

### Authors’ Contributions

Conceptualization, Pramil Singh and Anne Berit Petersen; Data analysis, Pramil Singh, Jerry Lee and Stacey Wiles; Funding acquisition, Anne Berit Petersen and Pramil Singh; Project administration, Anne Berit Petersen and Pramil Singh; Writing – original draft, Stacey Wiles; Writing – final draft, Pramil Singh, Anne Berit Petersen; Writing - review & editing, Jerry Lee, Pramil Singh, Anne Berit Petersen, and Anna Nelson.

## Data Availability

All data produced in the present study are available upon reasonable request to the authors.

## Author Acknowledgements

The authors wish to thank T. Allen Merritt, Gretchen Page, Maribel Munoz, Eva Arambula, and Diana Shouman-Garcia for their work on the Comprehensive Tobacco Treatment Program, as well as First 5CA.gov for funding the program.

## Notes

### Competing Interest Statement

The authors have declared no competing interest.

### Author Declarations

Loma Linda University Health Institutional Review Board granted approval to analyze individually de-identified data (No. 5190418).

